# Variant landscape of the *RYR1 gene* based on whole genome sequencing of the Singaporean population

**DOI:** 10.1101/2021.08.11.21261947

**Authors:** Claribel Tian Yu Foo, Yi Hui To, Astrid Irwanto, Alvin Yu-Jin Ng, Benedict Yan, Sophia Tsong Huey Chew, Jianjun Liu, Lian Kah Ti

## Abstract

**Background:** The *RYR1* gene codes for a ryanodine receptor which is a calcium release channel in the skeletal muscle sarcoplasmic reticulum. It is associated with Malignant Hyperthermia (MH) and several congenital myopathies including Central Core Disease (CCD), Multiminicore Disease (MMD) and Congenital Fibre-Type Disproportion (CFTD). There is currently little information on the epidemiology of *RYR1* variants in Asians. Our study aims to describe the *RYR1* variant landscape in a Singapore cohort unselected for *RYR1*-associated conditions.

**Methods:** Data was retrieved from the SG10K pilot project, where whole genome sequencing was performed on volunteers unselected and undetermined for *RYR1*-associated conditions. Variants were classified based on pathogenicity using databases ClinVar and InterVar. Allele frequencies of pathogenic variants were compared between Chinese, Indians and Malays. Using databases ExAC, GnomAD and GenomeAsia 100k study, we further compared local allele frequencies to those in Europe, America and Asia.

**Results:** Majority of the *RYR1* variants were missense mutations. We identified four pathogenic and four likely pathogenic *RYR1* variants. All were related to the aforementioned *RYR1*-associated conditions. There were 6 carriers of *RYR1* pathogenic variants amongst 4810 individuals, corresponding to an allele frequency of 0.06%. The prevalence of pathogenic variants was the highest amongst Indians (4 in 1127 individuals). Majority of pathogenic and likely pathogenic mutations were missense and located in mutational hotspots. These variants also occurred at higher frequencies in Asians than globally.

**Conclusion:** This study describes the variant landscape of the *RYR1* gene in Singapore. This knowledge will facilitate local efforts in developing precision medicine.

## Background

The *RYR1* gene (NCBI 2020) codes for a ryanodine receptor found in the skeletal muscle and the receptor serves as a calcium release channel in the sarcoplasmic reticulum. Mutations in *RYR1* are strongly associated with Malignant Hyperthermia (MH) and congenital myopathies such as Multiminicore Disease (MMD), Central Core Disease (CCD), and Congenital Fibre-Type Disproportion (CFTD) (Todd et al. 2021).

MH is a pharmacogenetic disorder of skeletal muscle which may be inherited in an autosomal dominant fashion or arise due to *de novo* mutations in associated genes, most commonly *RYR1* (Miller et al. 2018; Rosenberg et al. 2015). MH-causative mutations in *RYR1* result in increased sensitivity of *RYR1* protein to activation (Riazi et al. 2018; Robinson et al. 2006; Treves et al. 2008). In the presence of anesthetic triggering agents, dysregulation of calcium transport results in excessive myoplasmic calcium accumulation. This manifests clinically as a hypermetabolic crisis when an MH-susceptible individual is exposed to a volatile anesthetic agent or depolarizing muscle relaxants. This syndrome is potentially lethal if left untreated. The clinical manifestations of MH include initial hypercarbia, tachypnea, generalised muscle rigidity, masseter muscle rigidity, as well as arrhythmias. Subsequently, hyperthermia and rhabdomyolysis may occur (Litman 2019).

Congenital myopathies are inherited neuromuscular disorders characterised by common histologic features seen on muscle biopsy, such as central cores, nemaline rods, multi-minicores and central nuclei. *RYR1* is associated with several such myopathies, such as CCD, certain subtypes of MMD, and CFTD. There is significant genetic, phenotypic and histopathological overlap between these myopathies, as well as considerable phenotypic variability. CCD is inherited in an autosomal dominant fashion and is characterised by hip girdle weakness with orthopaedic complications such as hip dislocation and scoliosis. MMD is inherited in an autosomal recessive fashion and has a variety of clinical phenotypes. The most common subtype leads to spinal rigidity, scoliosis and respiratory dysfunction, and is associated with mutations in the selenoprotein N gene. Other subtypes are associated with *RYR1* mutations and can be characterised by hip girdle weakness, external ophthalmoplegia and distal muscle atrophy. Various studies have suggested that these subtypes of MMD may exist on a clinco-pathologic continuum with CCD (Jungbluth 2007; Zhou et al. 2007). CFTD can be inherited in an autosomal dominant or recessive pattern and rarely in an X-linked pattern (MedGen). It is characterised by weakness in the proximal muscle groups close to the trunk, such as the shoulder and hips. It can also result in ophthalmoplegia, ptosis (droopy eyelids) and scoliosis. *RYR1* mutations account for 10-20% of CFTD (Litman et al. 2018).

Many of the *RYR1*-associated myopathies are linked to MH-susceptibility, and it is estimated that MH-susceptibility and myopathy co-occur in at least 30% of individuals with *RYR1* pathogenic variants. As a result, patients with *RYR1*-associated myopathies should be considered MH-susceptible unless proven negative by an MH contracture biopsy (Litman et al. 2018).

Genetic testing is a common and non-invasive diagnostic test for many of these conditions. Most of the genetic studies done on RYR1 mutations were performed on patients diagnosed with MH and other myopathies, and this is useful in identifying pathogenic variants. However, to estimate the true population prevalence of these mutations, studies need to be conducted in the general population. Our review of the existing literature revealed one such variant landscape study on the general population, however this was conducted on the American population made up of mainly Caucasians (Gonsalves et al. 2013). There is little information on the epidemiology of *RYR1* variants in the Asian population. Our study performed on the multiracial Singapore population will be useful in providing a snapshot of the spectrum of mutations in Asia.

Our study aims to describe the *RYR1* variant landscape in a Singaporean cohort unselected for *RYR1*-associated myopathies and MH. We seek to define the prevalence of pathogenic variants in 3 major ethnic groups in Singapore and compare it to existing databases. This will facilitate genetic screening of these conditions in the Singapore population.

## Materials and Methods

Data was retrieved from the SG10K pilot project, which is part of a national effort to eventually whole-genome sequence 10,000 Singaporeans. The aim of this study was to provide a snapshot of the genetic diversity in East Asia, Southeast Asia, and South Asia. This is possible given the ethnic diversity in Singapore, which consists mainly of Chinese (74.3%), Malays (13.4%) and Indians (9.1%). The ethnic clusters in Singapore were analysed using principal-components analysis and are further described in the SG10K pilot project. In this study, whole genome sequencing was performed on 4810 samples, including 2780 Chinese, 903 Malays and 1127 Indians. These volunteers include a mixture of healthy and diseased individuals with conditions such as heart and neurodegenerative conditions. Linkage disequilibrium (LD)-based joint calling and quality controls were performed to remove low-quality variants and population-based phasing to obtain haplotypes. After removing relatedness up to the third degree, a subset of 4,441 unrelated individuals were obtained. Further details on the methodology can be obtained from the SG10K paper (Wu et al. 2019).

ClinVar and InterVar were used to classify variants into “benign”, “likely benign”, “uncertain significance”, “likely pathogenic” and “pathogenic”. ClinVar is a database of clinical variations which collects data on genotype-phenotype relationships from various contributors. It aggregates the submissions and reports the clinical significance of variants based on the American College of Medical Genetics (ACMG) and Association for Molecular Pathology (AMP) Guidelines (NCBI). InterVar is a bioinformatics software which classifies variants via automation of the 2015 ACMG/AMP guidelines (Li and Wang). ClinVar serves as our main reference for determining the clinical significance of variants as the classifications are more thoroughly supported with published evidence. The variant positions for RYR1 refer to NM_001042723.

Variants were also classified based on the location and type of mutations. Single Nucleotide Variant (SNV) types include synonymous SNV, missense SNV, stop gain SNV, stop loss SNV and unknown. Indel variant types include frameshift insertion, frameshift deletion, stop gain, stop loss, non-frameshift insertion, non-frameshift deletion and unknown.

Allele frequencies were compared between the Chinese, Indian and Malay population in Singapore. Allele frequencies in the Singapore population were also compared against European, American and Asian populations using databases ExAC (Exome Aggregation Consortium) (ExAC), GnomAD (Genome Aggregation Database) (gnomAD), and data from the GenomeAsia 100k study (GenomeAsia).

These studies were approved by the Institutional Review Board of the National University of Singapore (Approvals: N-17-030E and H-17-049), SingHealth Centralized Institutional Review Board (Approvals: 2006/612/A, 2014/160/A, 2018/2570, 2018/2717, 2010/196/C, 2015/2194, 2019/2046, 2009/280/D, 2014/692/D, 2002/008/A, 2002/008 g/A, 2013/605/C, 2018/3081 and 2015/2308), National Health Group Domain Specific Review Board (Approvals: 2007/00167, 2016/00269, 2018/00301, TTSH/2014-00040 and 2009/00021).

Statistical analysis was not performed as the number of variants is small and significance testing is not meaningful.

## Results

### Types of mutations found

All the *RYR1* variants were classified based on mutation type (Figure 1). All of these mutations occurred in exonic regions. Majority of the mutations were missense, followed by synonymous variants.

**Fig. 1.**
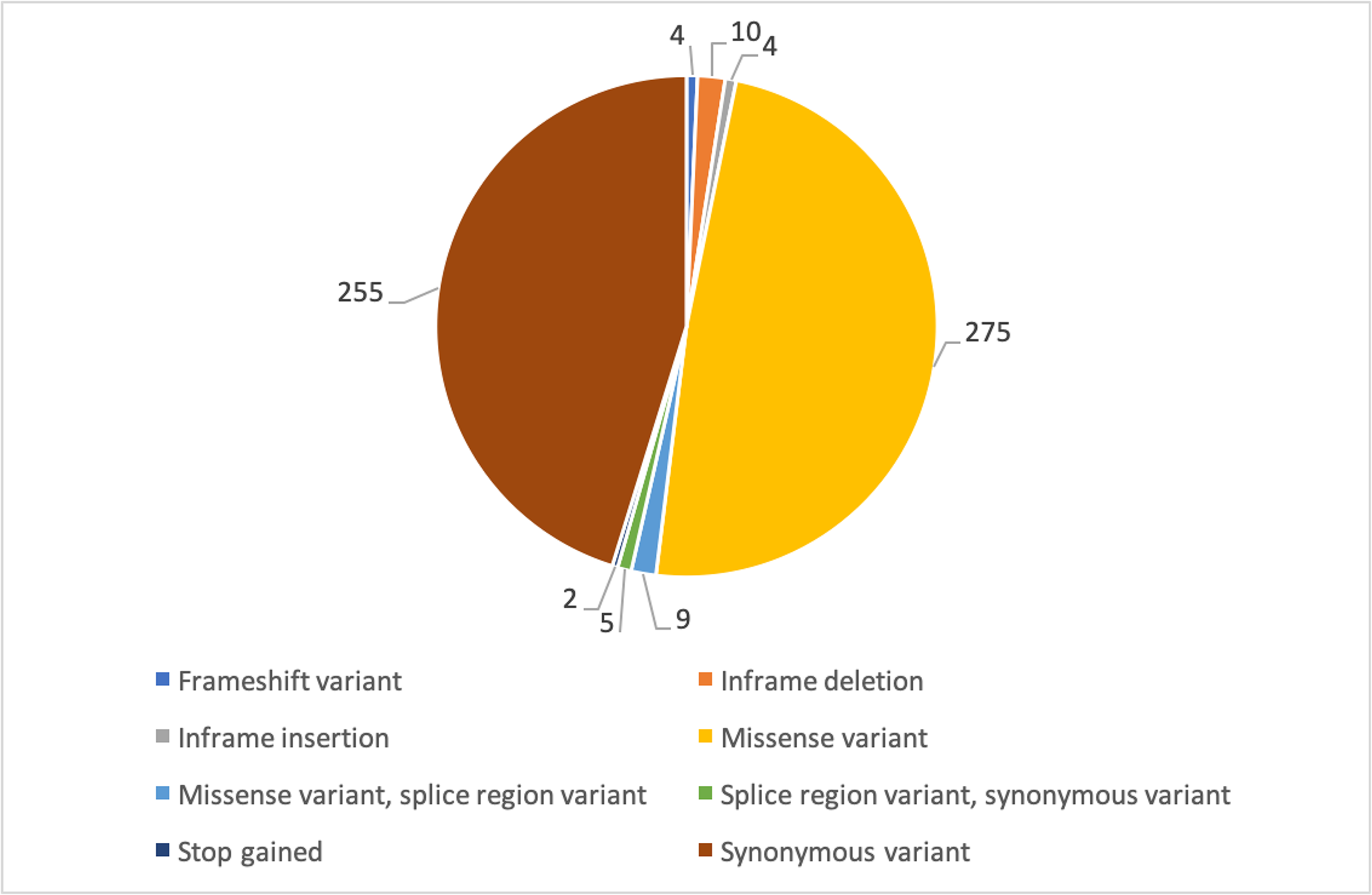
Mutation counts and types for *RYR1* variants in SG10K pilot data

### Classification of *RYR1* variants based on clinical significance

The *RYR1* variants found in our study were classified based on their clinical significance given by ClinVar (online resource table S1) and InterVar (online resource table S2). ClinVar was used as the main reference for defining the clinical significance of the variants. Based on ClinVar, there were 4 pathogenic and 2 likely pathogenic variants for *RYR1*. Based on InterVar, there were 1 pathogenic and 6 likely pathogenic *RYR1* variants. Of these, all overlapped with the pathogenic and likely pathogenic variants found in ClinVar except for 2 likely pathogenic variants. Table 1 summarizes the list of pathogenic and likely pathogenic *RYR1* variants found in our study. Table 2 and online resource table S3 provide details on the specific diseases associated with each variant. The majority of pathogenic and likely pathogenic *RYR1* variants in our study were missense mutations, except for 2 nonsense mutations (Table 1).

**Table 1:** Pathogenic and likely pathogenic *RYR1* variants in the Singapore population

| Variant | ClinVar classification | InterVar classification | Number of carriers | Ethnicity of carriers | Exon number | Type of mutation |
| --- | --- | --- | --- | --- | --- | --- |
| c.325C>T<br>(p.Arg109Trp) | Pathogenic | Likely pathogenic | 1 | Chinese | 4/106 | Missense |
| c.6487C>T<br>(p.Arg2163Cys) | Pathogenic | Likely pathogenic | 1 | Malay | 39/106 | Missense |
| c.7268T>A<br>(p.Met2423Lys) | Pathogenic | Variant of uncertain significance | 3 | Indian | 45/106 | Missense |
| c.14111C>T<br>(p.Thr4704Met) | Pathogenic | Likely pathogenic | 1 | Indian | 96/106 | Missense |
| c.742G>T<br>(p.Gly248Trp) | Likely pathogenic | Likely pathogenic | 1 | Chinese | 12/106 | Missense |
| c.1186G>T<br>(p.Glu396X) | NA* | Pathogenic | 3 | Malay | 9/106 | Stop gained |
| c.2654G>A<br>(p.Arg885His) | Conflicting** | Likely pathogenic | 3 | Indian | 21/106<br>(outside mutational hotspot) | Missense |
| c.8554C>T<br>(p.Arg2852X) | Variant of uncertain significance | Likely pathogenic | 1 | Chinese | 55/106<br>(outside mutational hotspot) | Stop gained |

**Table 2:**
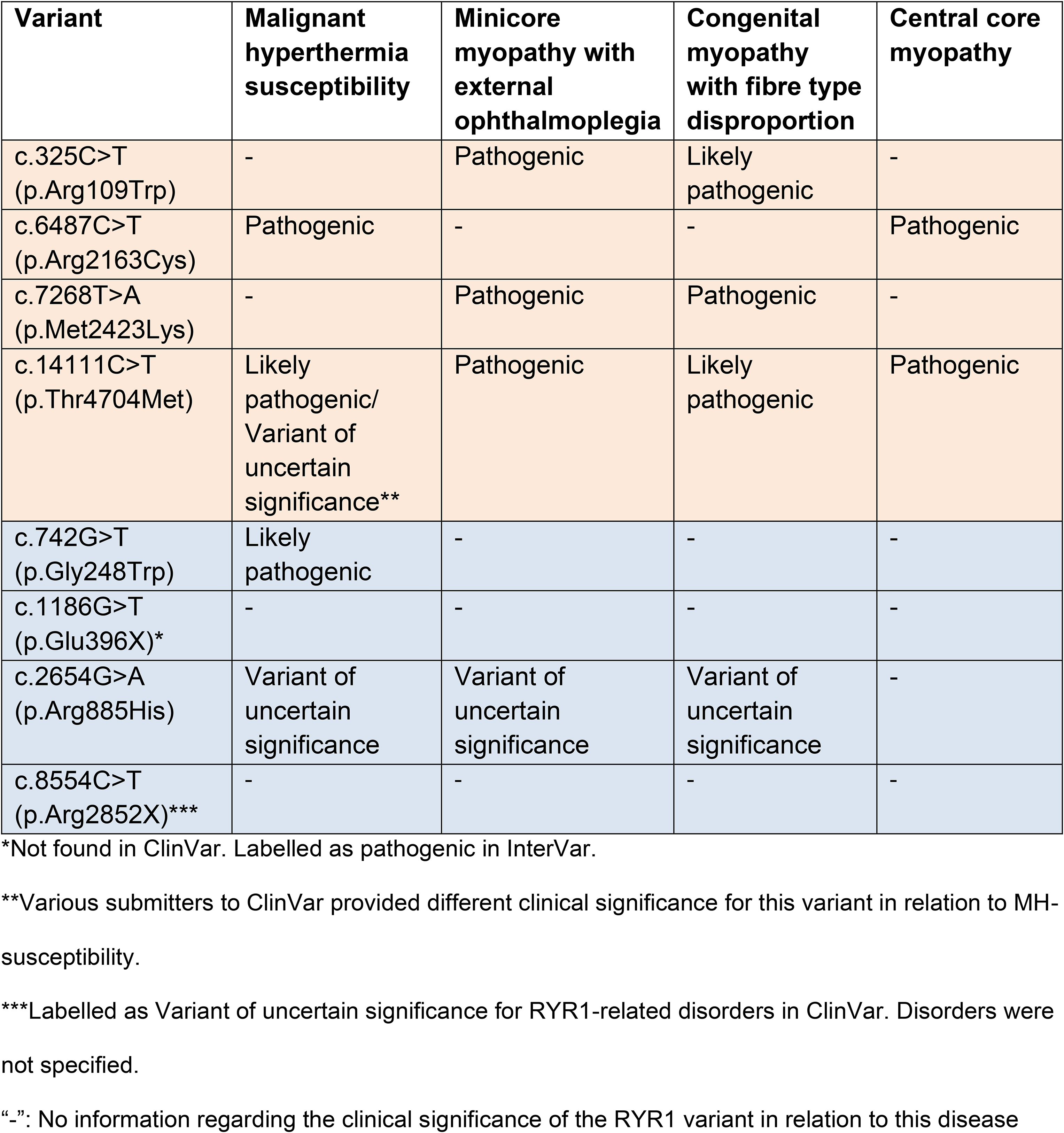
Diseases associated with pathogenic and likely pathogenic *RYR1* variants

In terms of the location of pathogenic and likely pathogenic *RYR1* mutations, all but 2 likely pathogenic variants were found in mutational hotspots. These *RYR1* mutational hotspots have been previously identified to include the N-terminal region between codons 34 and 614 (exons 2 to 19), central region between codons 2163 and 2458 (exons 39 to 47), and C-terminal region between codons 4136 and 4973 (exons 85 to 104) (Gillies et al. 2008). There was no clear pattern as to the exons in which the pathogenic and likely pathogenic mutations were found. However, we found that most of the pathogenic and likely pathogenic *RYR1* variants have mutations occurring in highly conserved regions (online resource table S4).

### Ethnic distribution of the carriers of the pathogenic *RYR1* variants

The ethnic distribution of pathogenic *RYR1* variants was analysed. Indians made up the majority (67%) of the individuals who carried pathogenic variants (online resource figure S1).

On further analysis of the ethnic distribution, each *RYR1* variant was found exclusively in a single ethnicity. Of the 4 pathogenic variants identified by ClinVar, 1 was present in Chinese, 2 in Indians, and 1 in a Malay. All variants were present at similar frequencies (only 1 carrier), except c.7268T>A which was present in 3 Indian carriers (Figure 2).

**Fig. 2.**
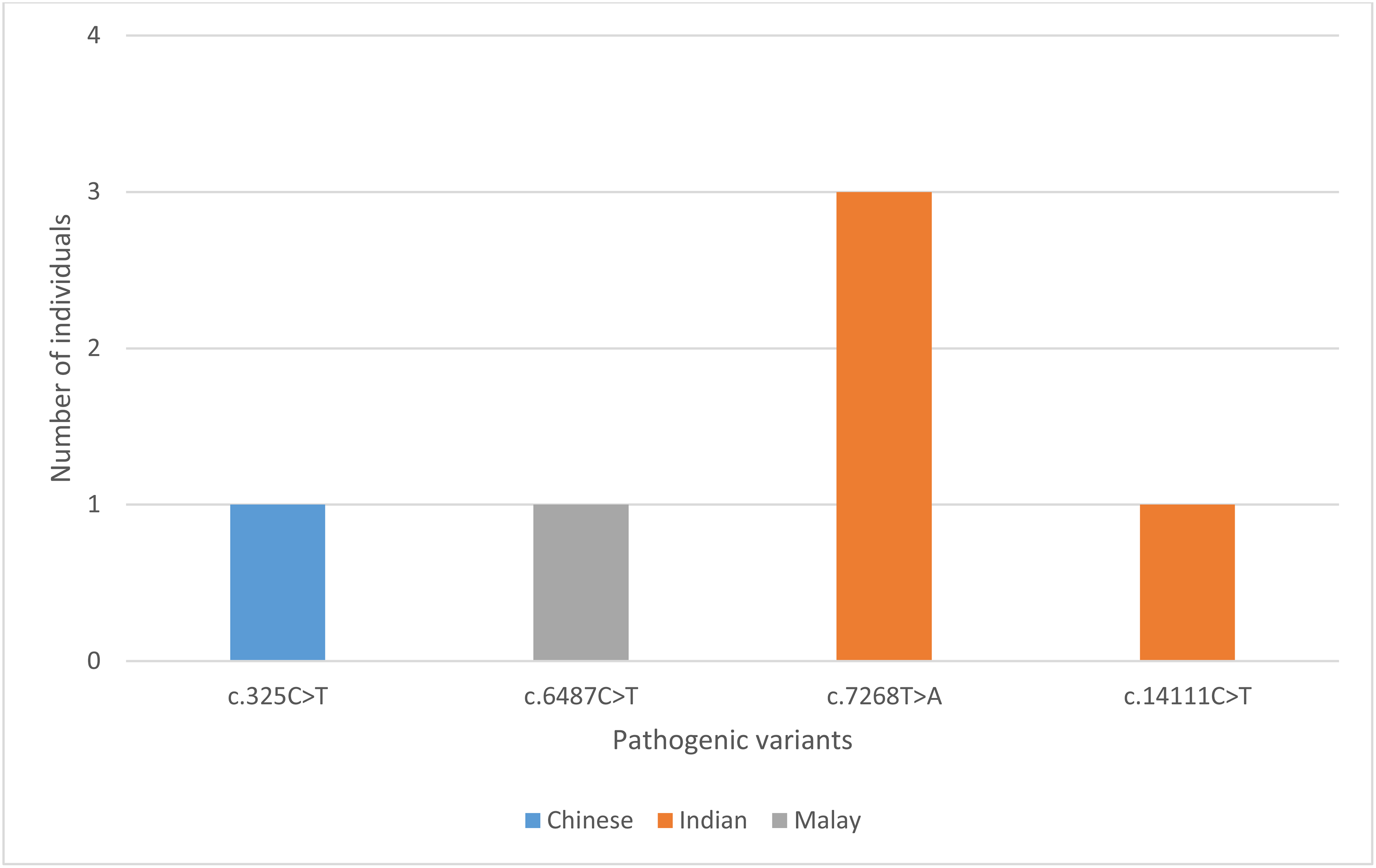
Ethnic distribution of individual pathogenic *RYR1* variants

The analysis was repeated with both pathogenic and likely pathogenic *RYR1* variants. Indians still made up the largest proportion of carriers (50%) (online resource figure S2). Further analysis similarly revealed that each *RYR1* variant was unique to each ethnicity. Of the 8 pathogenic and likely pathogenic variants, 3 were present in the Chinese, 2 were present in the Malays and 3 in the Indians. Three of the variants occurred at higher frequencies – c.7268T>A was found in 3 Indians as previously mentioned, c.1186G>T was found in 3 Malays and c.2654G>A in 3 Indians (online resource figure S3).

### Allele frequencies across different populations

Allele frequencies of 2 of the pathogenic *RYR1* variants (c.7268T>A and c.14111C>T) found in our study were higher in Singapore than globally (online resource table S5). c.325C>T occurred at higher frequencies in the Non-Finnish European population based on the ExAC database. c.6487C>T occurred at higher frequencies in the South Asian population based on the gnomAD database. Amongst the likely pathogenic variants, 3 variants (c.1186G>T, c.2654G>A and c.8554C>T) occurred at higher allele frequencies in Singapore than globally. Only variant c.742G>T had a higher allele frequency in the South Asian population based on the GenomeAsia 100K study.

In sum, allele frequencies of the pathogenic and likely pathogenic variants found in our study were generally higher amongst Asians (Singapore population and South Asian population) than globally. Comparing between the Singapore population and South Asian population, different databases have provided different allele frequencies for the South Asian population and therefore we are unable to conclude if these variants occur at higher frequencies in either population. Data from other parts of Asia is also lacking.

## Discussion

Most of the genetic studies on *RYR1* have been conducted on populations selected for *RYR1*-associated disorders, and there is limited information on the epidemiology of *RYR1* variants in the Asian population. This is the first paper that analyses the genetic landscape of *RYR1* specifically in the Singapore population.

The prevalence of pathogenic *RYR1* variants in our study was 6 in 9620, corresponding to an estimate of 1 in 2000 in the Singapore population. Interestingly, we found that the prevalence of pathogenic *RYR1* variants in Indians was 4 in 1127, much higher than in Chinese and Malays (1 in 2780 and 1 in 903 respectively). The allele frequencies of the pathogenic variants found in our study were generally higher in Asians (Singapore population and South Asian population) than globally. In terms of the location of *RYR1* mutations, those found in our study largely occurred within established mutational hotspots.

### Significance of the pathogenic variants found

The pathogenic *RYR1* variants found were closely associated with diseases such as MH, MMD, CCD and CFTD.

*RYR1* gene encodes the Ca^2+^-dependent, Ca^2+^ channel ryanodine receptor 1 (*RYR1*) found on the sarcoplasmic reticulum of the skeletal muscle (Beam et al. 2016).

MH occurs due to dysregulation of Ca^2+^ transport caused by abnormal functioning of the skeletal muscle excitation-contraction (EC) coupling complex which comprises of an L-type voltage-gated Ca^2+^ channel (dihydropyridine receptor, DHPR) and a Ca^2+^-dependent, Ca^2+^ channel (*RYR1*). Excessive Ca^2+^ release from the sarcoplasmic reticulum causes uncontrolled muscle contraction (Santulli et al. 2018).

Mutations in the *RYR1* gene are also a common cause of congenital myopathies. Dominant mutations have been associated with CCD, while recessive mutations are associated with MMD and CFTD. Mutations associated with CCD leads to chronic channel dysfunction either through excitation-contraction uncoupling or persistent channel leakiness. However, little is known about recessive mutations and their mechanism of illness (Amburgey et al. 2013).

Four pathogenic *RYR1* variants were found and depicted in Figure 3. The N-terminal domains (NTD) make key interactions with other domains to stabilise the cytosolic assembly, and these interactions have also been implicated in channel gating and pore opening. c.325C>T is located in NTD-A, and both c.6487C>T and c.7268T>A are located in the helical domain (B-Sol). Both NTD-A and B-Sol are involved in key interprotomer contacts (Wang et al. 2005). In the context of MH, mutations at these points of contact may weaken the interfaces and facilitate pore opening and spurious Ca^2+^ leak (Santulli et al. 2018). c.6487C>T is a known causative mutation for MH and the amino acid substitution alters the conformational state of *RYR1*, exposing the receptor’s activating sites to stimulatory agents (Treves et al. 2005). c.325C>T and c.7268T>A occur due to mutations affecting highly conserved amino acid residues and are known to be autosomal recessive in relation to *RYR1*-related myopathies (Zhiguang Yuchi 2013; Zhou et al. 2007). However, they have yet to be studied in relation to the MH-susceptibility phenotype.

**Fig. 3.**
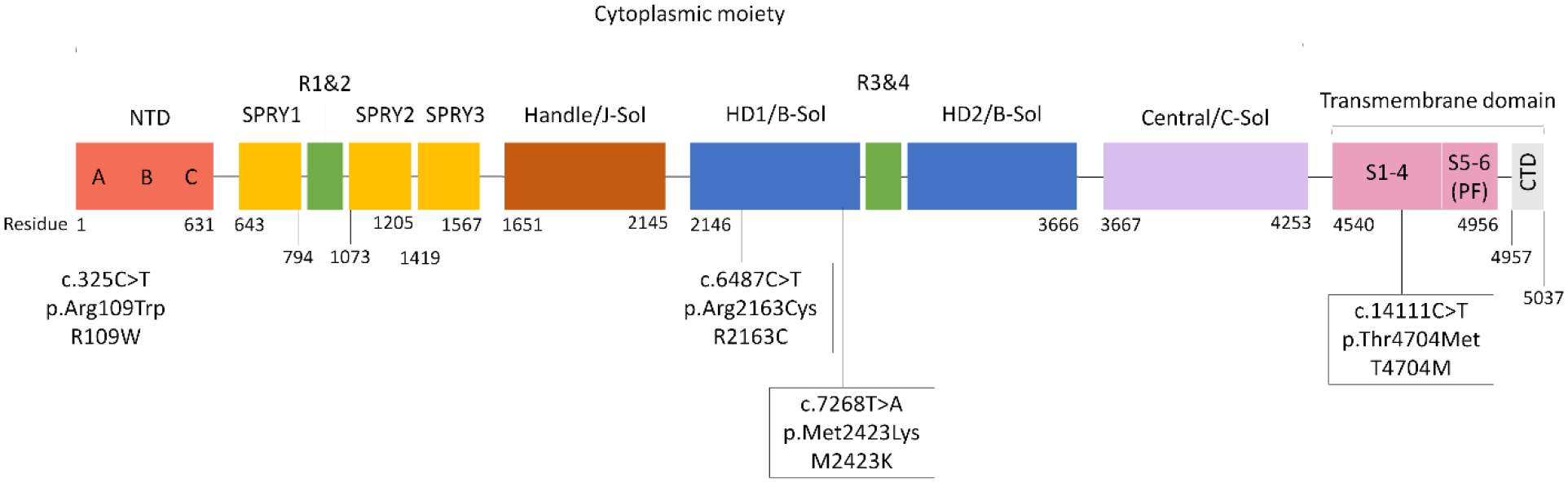
Schematic diagram of *RYR1* protein subunit and pathogenic variants in this study *RYR1* is a homotetramer with each subunit consisting of a large cytoplasmic moiety and a transmembrane domain. This figure depicts one subunit of the *RYR1* protein. The cytoplasmic moiety contains (1) the N-terminal domain (NTD) with 3 subdomains (A, B, C), (2) 3 SPRY domains (SPRY 1, 2, 3), of which SPRY 2 and 3 interact with the α1 subunit of DHPR at the II-III loop region, (3) a solenoid region comprising the handle/ junctional solenoid (J-sol) domain, and the helical (HD1, HD2)/ bridging solenoid (B-Sol) domains, (4) 2 tandem repeat domains (R1&2, R3&4), and (5) a Central domain or core solenoid (C-Sol). The transmembrane domain consists of 6 transmembrane helices (S1-6), and a C-terminal domain (CTD). S5 and 6, together with a pore helix in between S5 and S6, make up the pore-forming region (PF) [19, 25, 36]. The amino acid residue numbers for the various domains have been labelled, and the pathogenic *RYR1* mutations are indicated at their various positions across the protein

c.14111C>T affects a highly conserved amino acid residue (Zhou et al. 2007) located within the transmembrane helices S2-S3. It is hypothesised that this region participates in calcium-sensing and calcium-dependent inactivation of *RYR* (Zalk et al. 2015). This mutation is known to be autosomal recessive in relation to *RYR1*-related myopathies (Zhou et al. 2007). There is conflicting evidence for the clinical significance of this *RYR1* variant in relation to MH, with some suggesting that it is likely pathogenic for MH and others suggesting that it is a variant of unknown significance (NCBI 2019). Further studies are needed to definitively conclude its relation to MH-susceptibility.

### Limitations

This study is a retrospective study done using anonymised genetic data from the SG10K study. We therefore lacked access to the medical histories of the study participants. Hence, we are unable to determine if the carriers of these pathogenic *RYR1* variants displayed phenotypes of the associated diseases, and we also cannot determine the penetrance of these mutations.

As we have mentioned, our study population contains a mixture of healthy and diseased individuals with comorbidities such as eye, neurodegenerative, metabolic, cardiac diseases. However, *RYR1* is not found to be associated with these diseases. Hence the enrichment of mutations within *RYR1* should not be related to the presence of such co-morbidities within the sample population.

### Application

According to the EMHG guidelines, evaluation of patients for MH-susceptibility involves assessing for risk factors such as family history of MH or unexplained perioperative death, personal adverse reaction to general anaesthesia, postoperative or exertional rhabdomyolysis or exertional heat stroke. For patients with such risk factors, further testing is recommended. The decision between IVCT and genetic testing depends on several factors: the availability of the respective test, the urgency of test, the prior probability of a positive diagnosis and the cost of the tests. Genetic testing offers a less invasive option but is less sensitive than IVCT. Patients with a negative genetic screening result may proceed to do an IVCT to conclusively exclude MH susceptibility. Genetic testing has a major role in family screening and its cost-effectiveness makes it a viable option as the primary investigation of index cases (Girard 2015). The North American MH Genetic Group also adopts a similar approach by incorporating genetic testing into its screening protocol (Nelson et al. 2004). Disclosure of incidental findings from MH-susceptibility genetic screening in generally healthy individuals has also been found to reduce cost by USD $900 and increase quality-adjusted life years (QALYs) by 0.09 (Bennette et al. 2015).

Currently there is no active screening for MH-susceptibility in Singapore due to the fact that MH is a rare condition and that genetic testing is still relatively costly to the individual. Screening for malignant hyperthermia is only done through history-taking. Genetic testing will only be offered for individuals with clinical suspicion of MH-susceptibility. Currently, muscle contracture test is not available in Singapore. However, as Singapore moves towards precision medicine with initiatives such as the National Precision Medicine Initiative, the findings in our study will be valuable in helping to expanding our knowledge of the variant landscape of various genes in Singapore. This will facilitate subsequent efforts to provide cost effective genetic testing and personalised medicine to patients in the future.

Since there is significant overlap between MH susceptibility and congenital myopathies, we recommend genetic screening for malignant hyperthermia in patients with congenital myopathies and vice versa (Litman et al. 2018).

## Conclusion

From this study, we conclude that the prevalence of the pathogenic *RYR1* variants found in our study is higher amongst Asians than globally. Indians made up the majority of the individuals who carried pathogenic variants, compared to Chinese and Malays. The pathogenic variants were associated with diseases such as MH, MMD, CCD and CFTD. This paper also confirms that the variants found in our population are indeed rare and unique to individuals, suggesting a need to conduct genetic screening using sequencing rather than methods which look at specific mutations known to the database.

## Supporting information

Online resource figure S1

Online resource figure S2

Online resource figure S3

Online resource table S1

Online resource table S2

Online resource table S3

Online resource table S4

Online resource table S5

## Data Availability

The accession number for the sequence data reported in this paper is EGA: EGAS00001003875.

## Acknowledgments

We thank all the individuals who have contributed to this study.

## Abbreviations

MH: malignant hyperthermia
SNV: single nucleotide variants

## Declarations

### Availability of data and material

The accession number for the sequence data reported in this paper is EGA: EGAS00001003875.

### Code availability

The accession number for the sequence data reported in this paper is EGA: EGAS00001003875.

### Consent to participate

This study was conducted retrospectively using data obtained from the SG10K pilot project. Written informed consent was obtained from all participants of the SG10K pilot project.

### Consent for publication

This study was conducted retrospectively using data obtained from the SG10K pilot project. Written informed consent was obtained from all participants of the SG10K pilot project.

## List of supplemental material

**Online resource figure S1:**
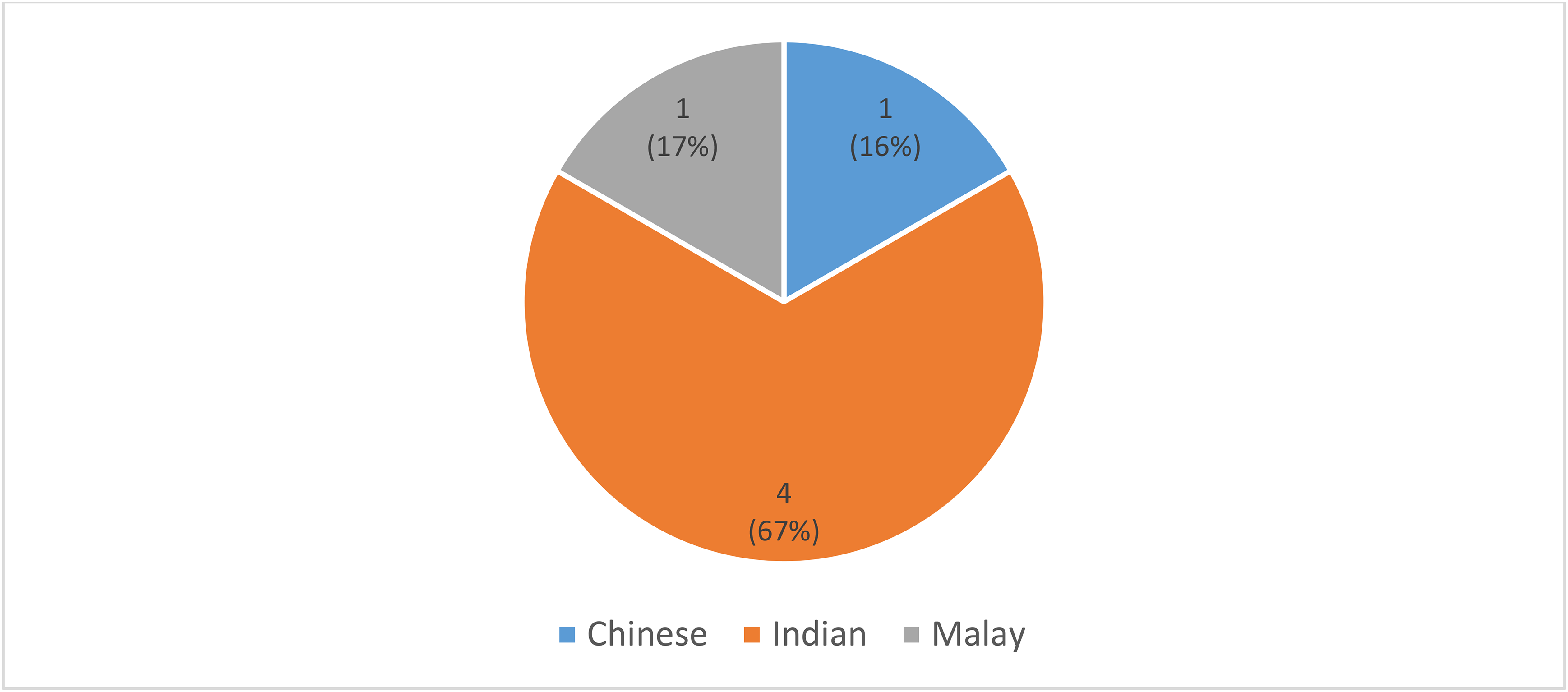
Ethnic distribution of all pathogenic *RYR1* variants

**Online resource figure S2:**
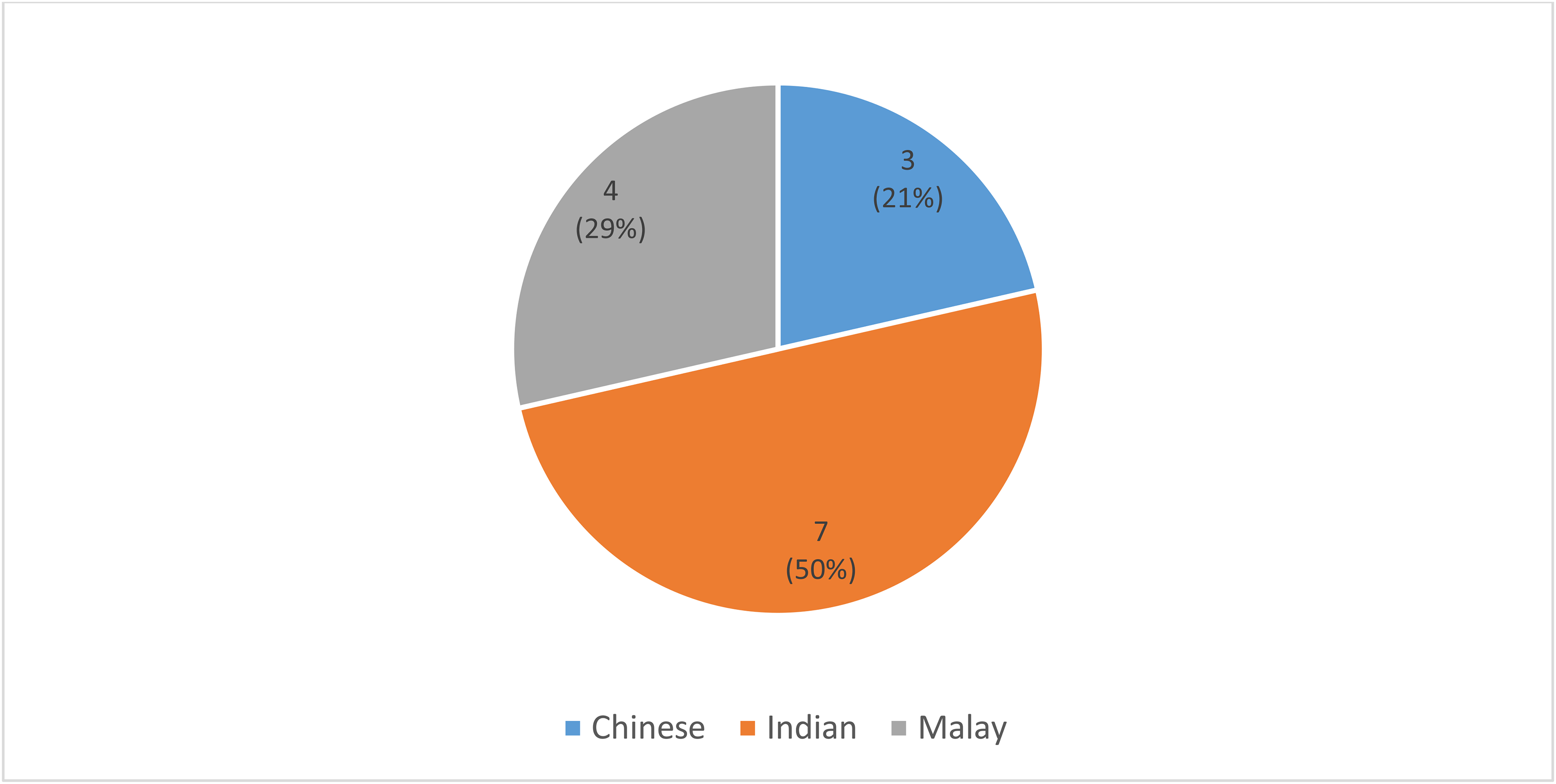
Ethnic distribution of all pathogenic and likely pathogenic *RYR1* variants

**Online resource figure S3:**
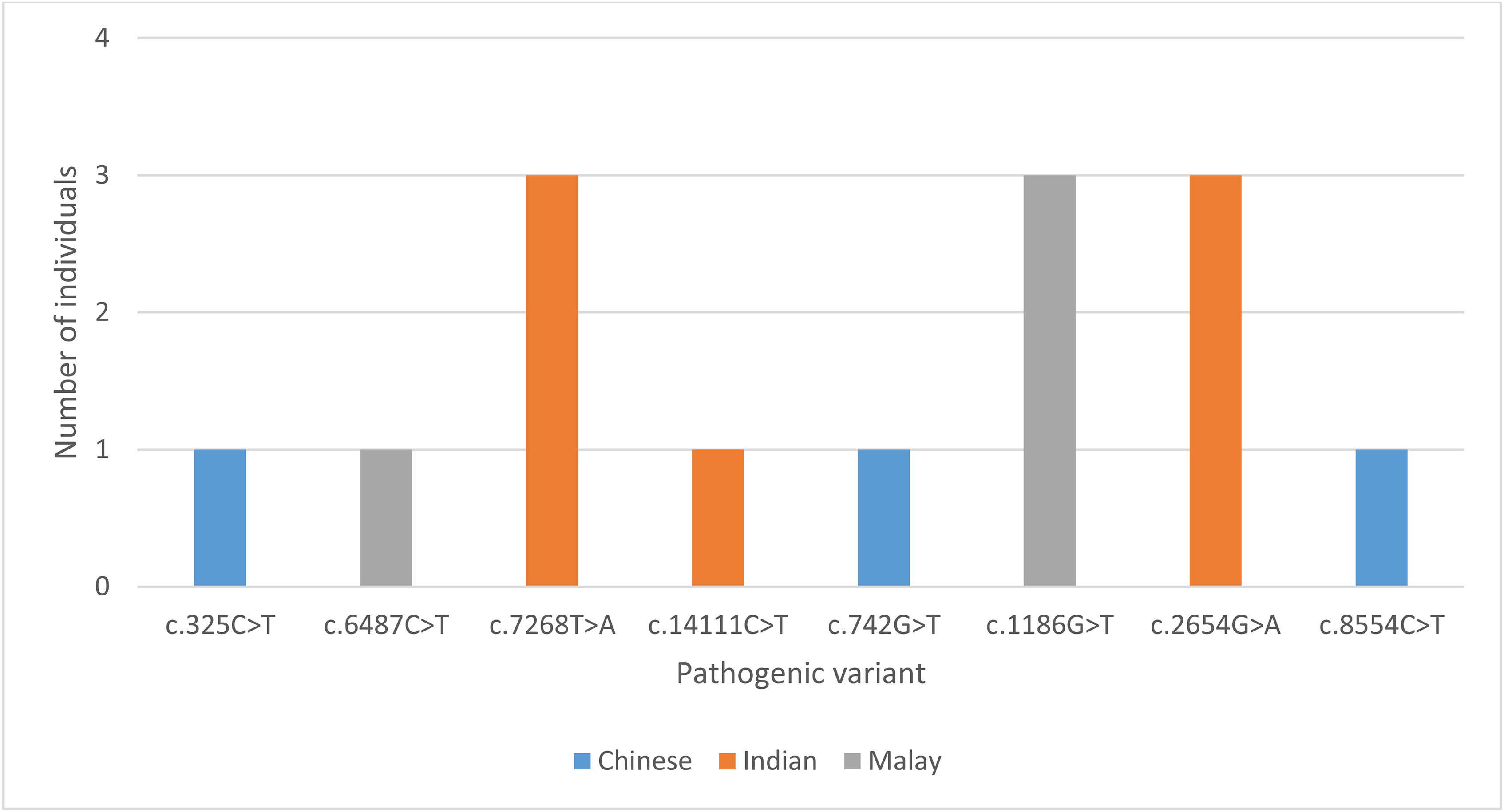
Ethnic distribution of individual pathogenic and likely pathogenic *RYR1* variants

**Online resource table S1:**
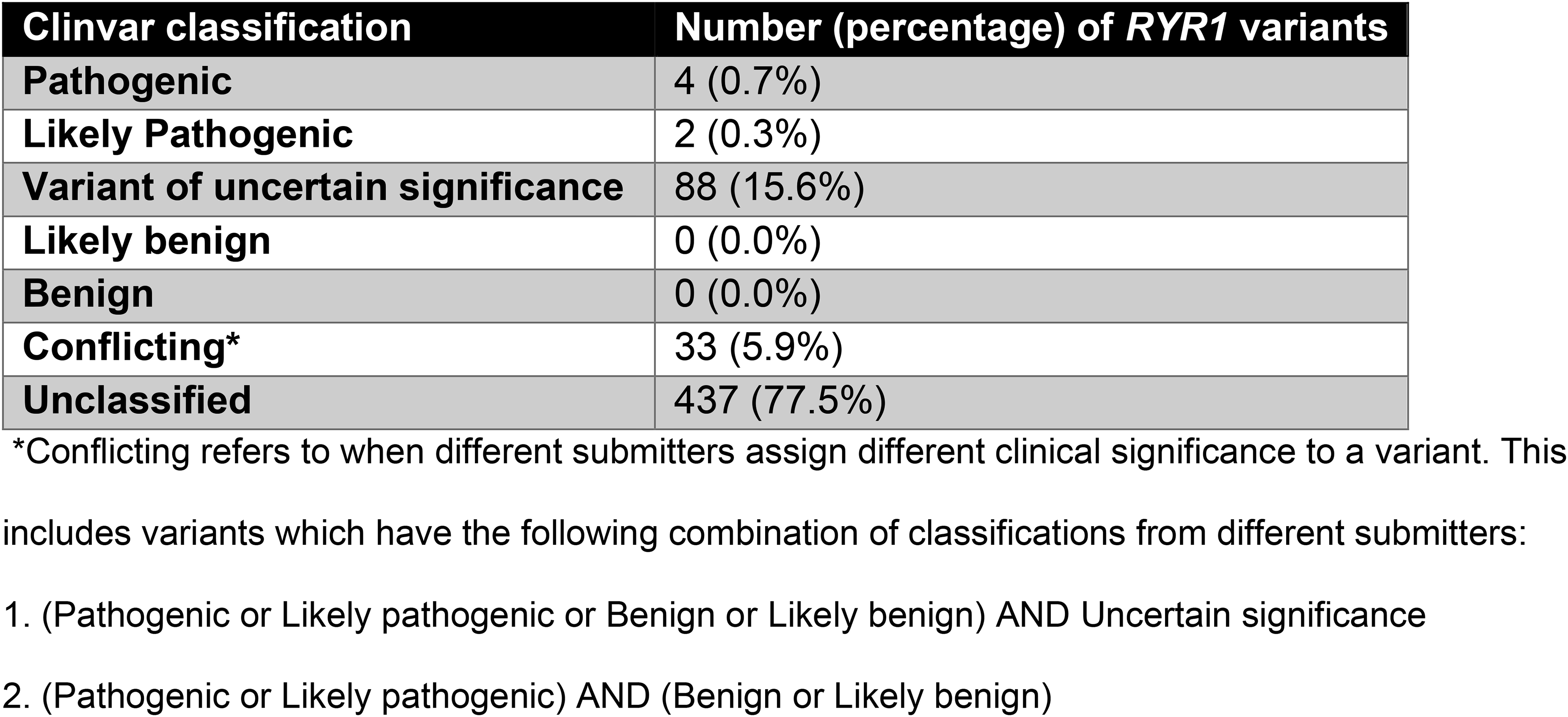
Clinical significance of *RYR1* variants based on ClinVar.

**Online resource table S2:**
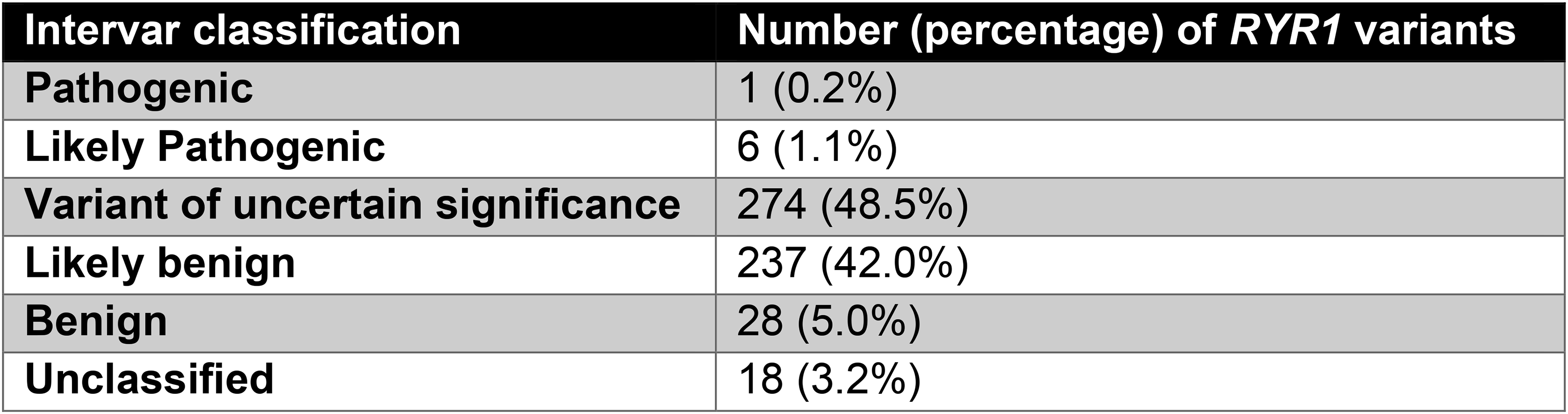
Clinical significance of *RYR1* variants based on InterVar.

**Online resource table S3:**
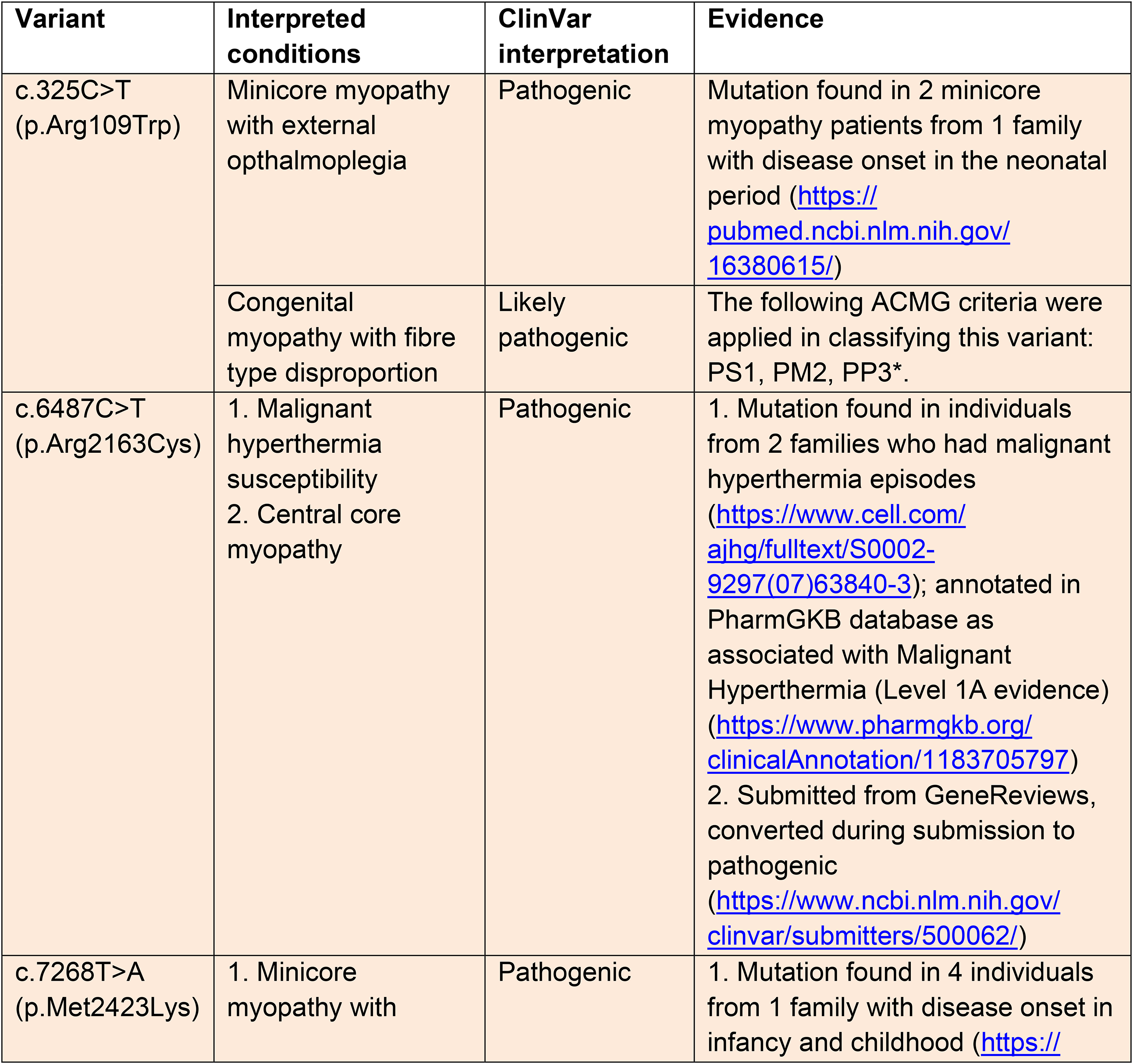
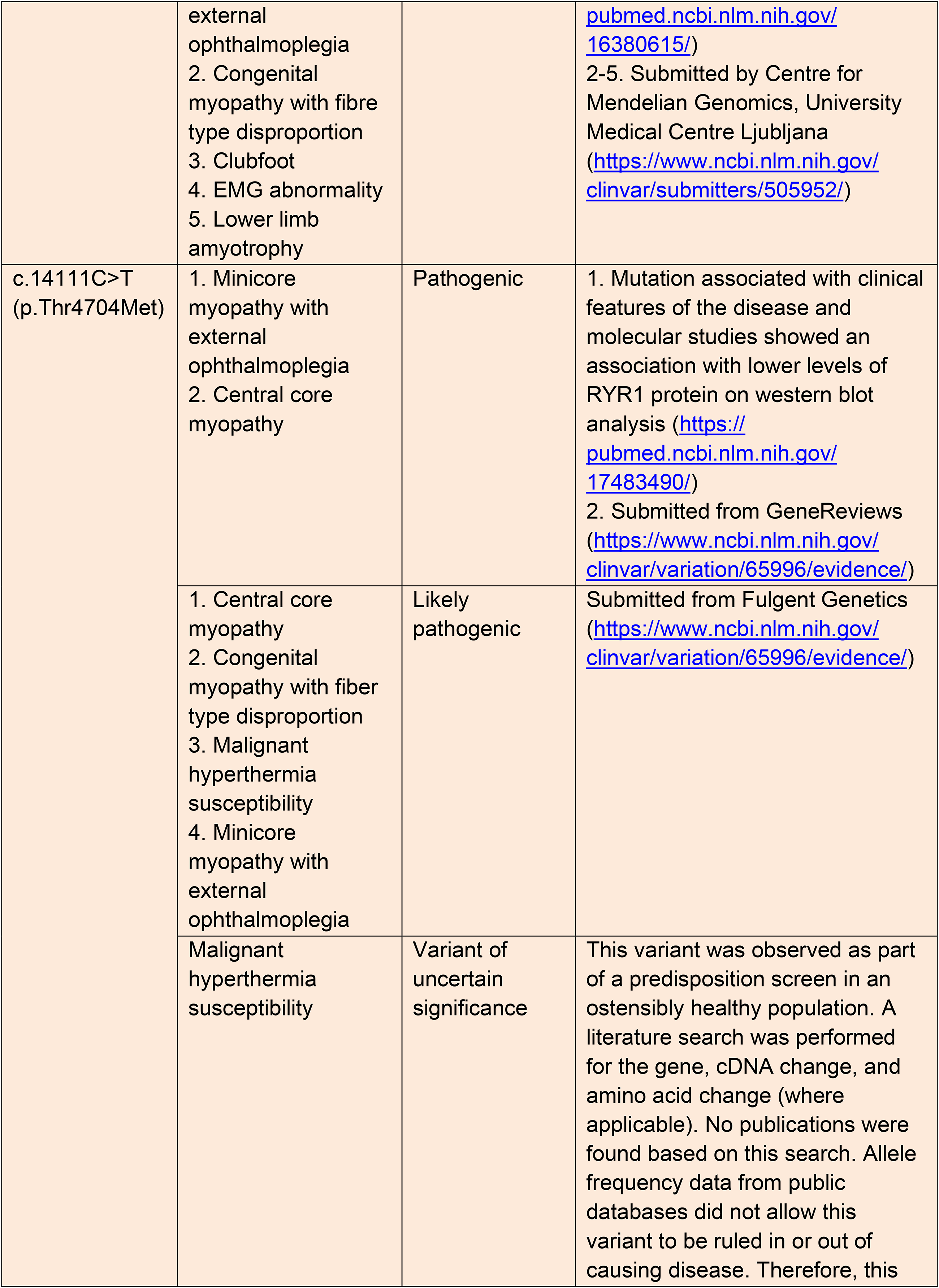
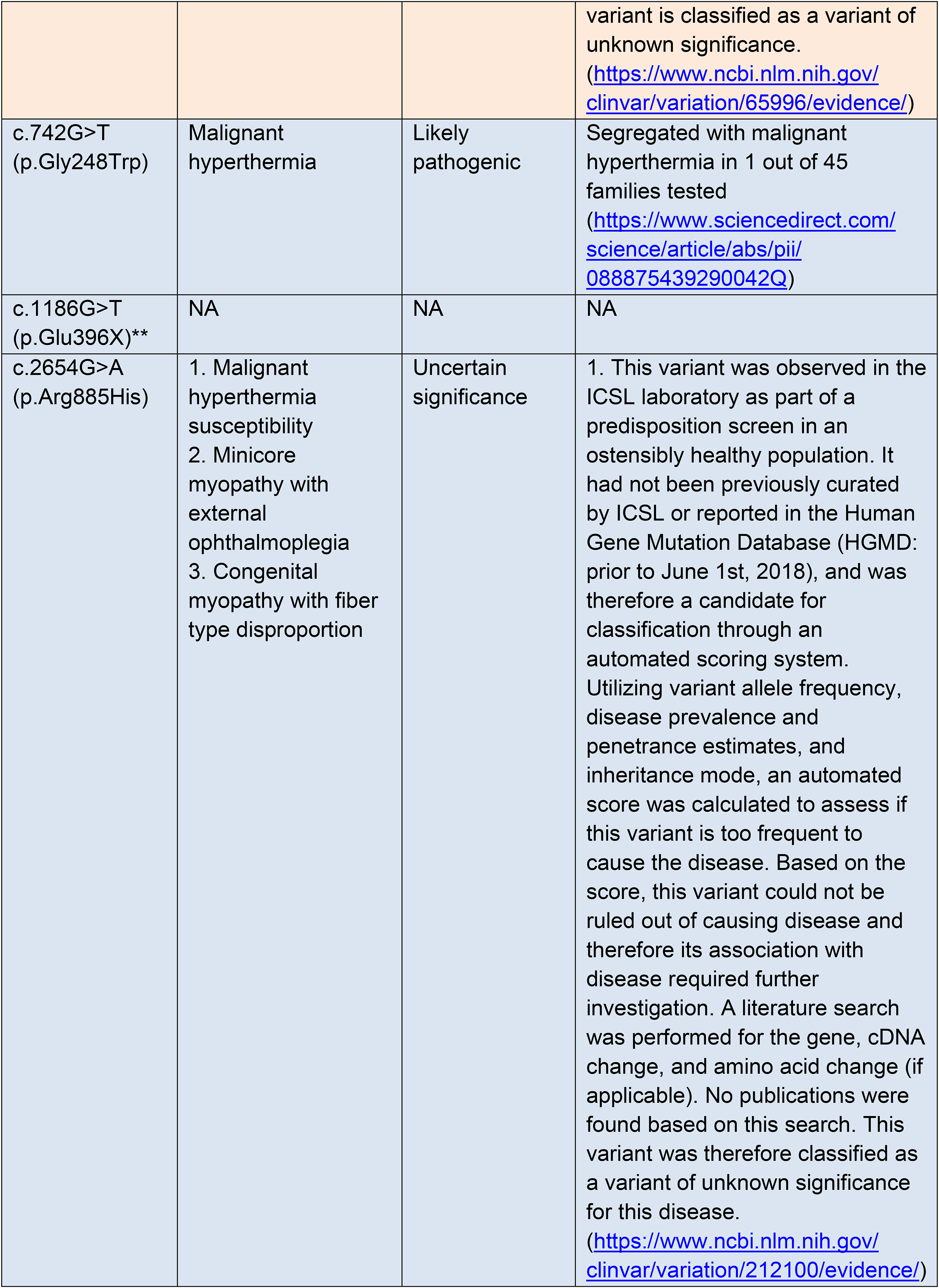
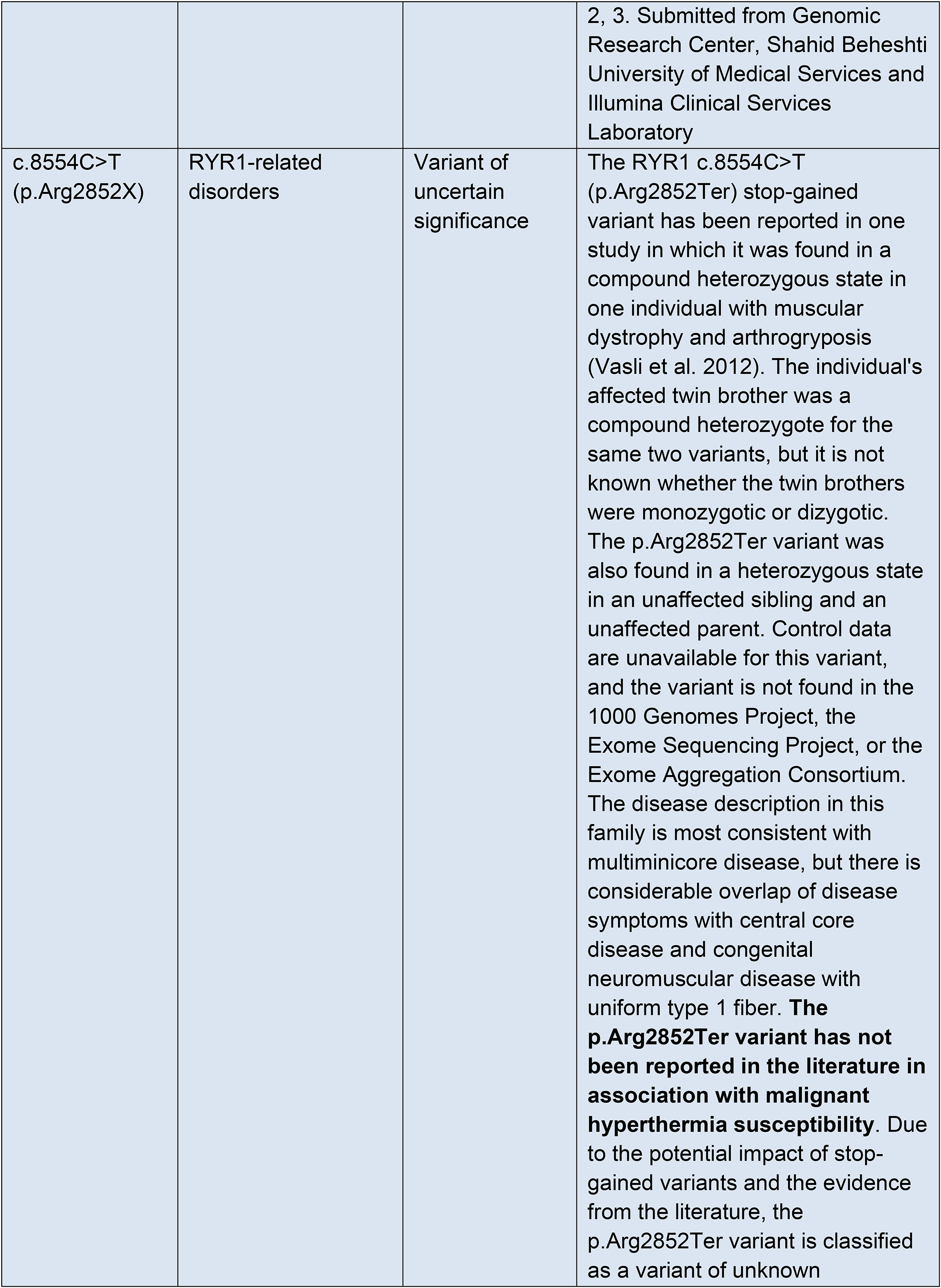
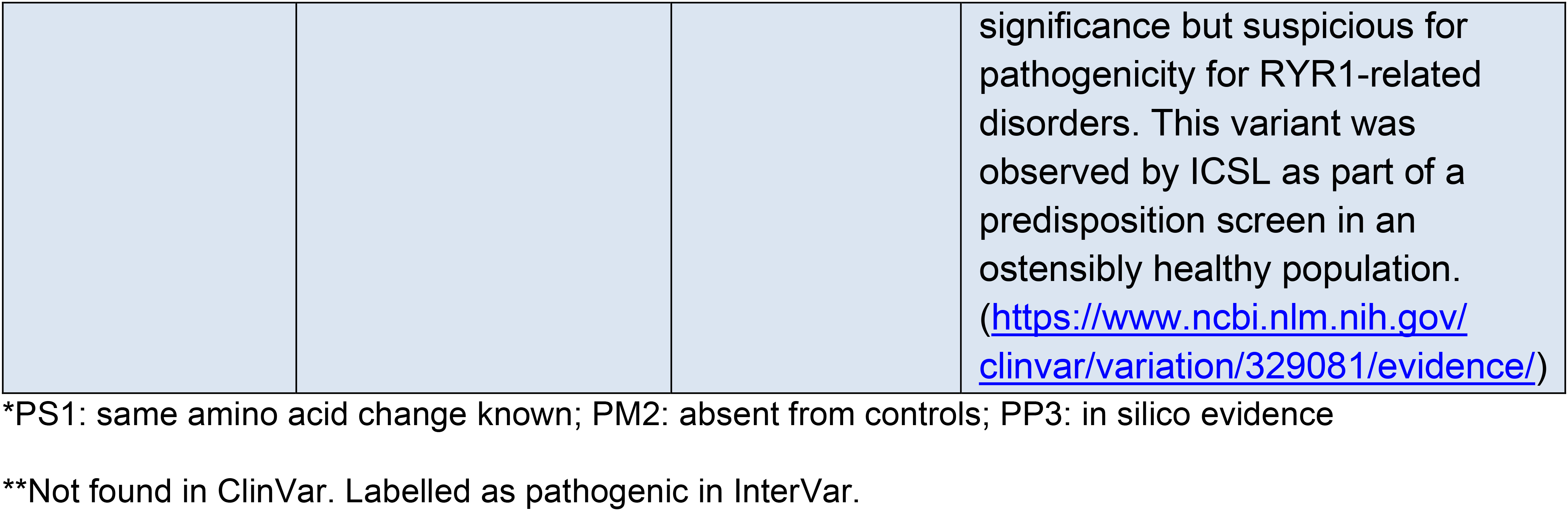
Diseases associated with pathogenic and likely pathogenic RYR1 variants with supporting evidence from ClinVar

**Online resource table S4:**
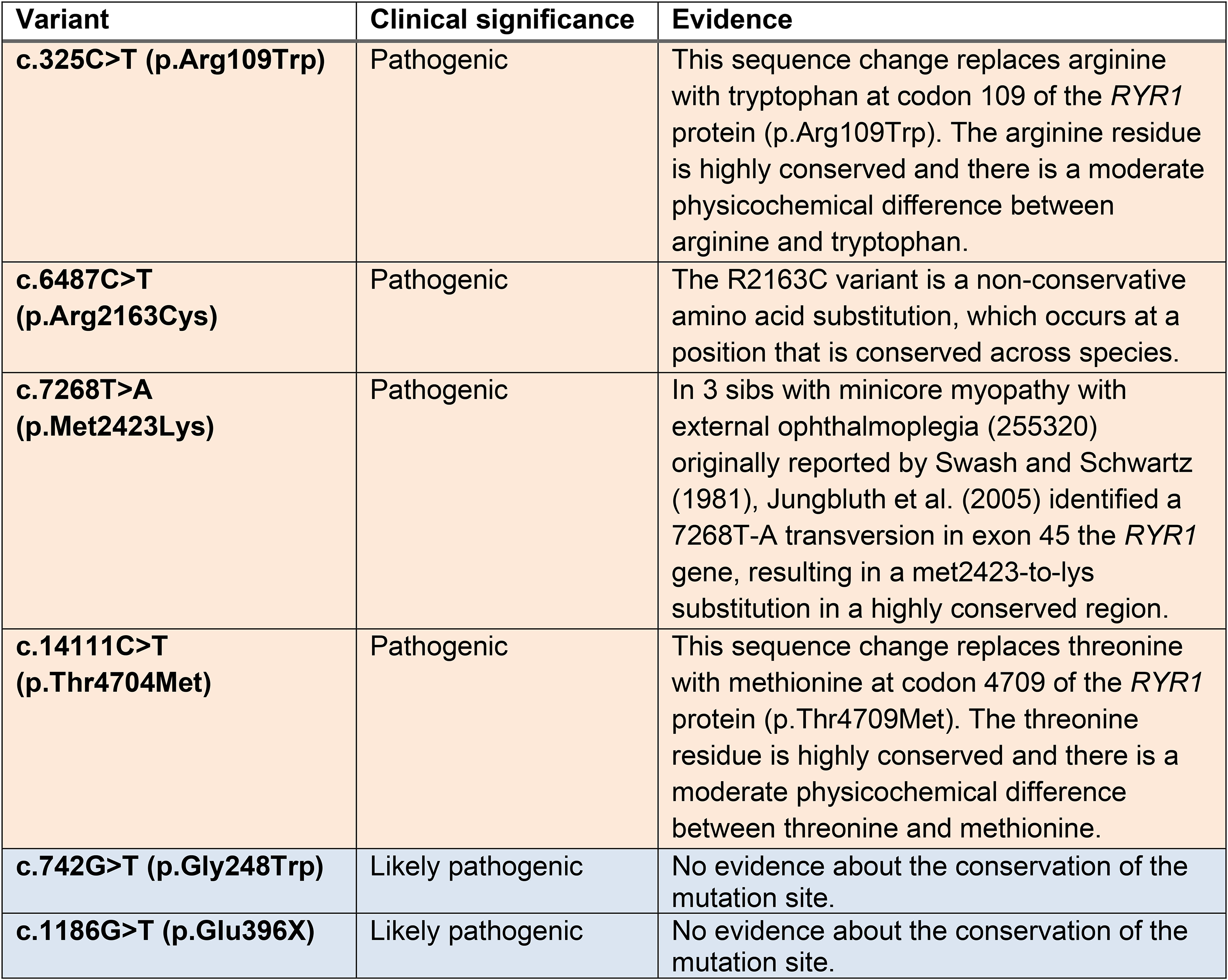
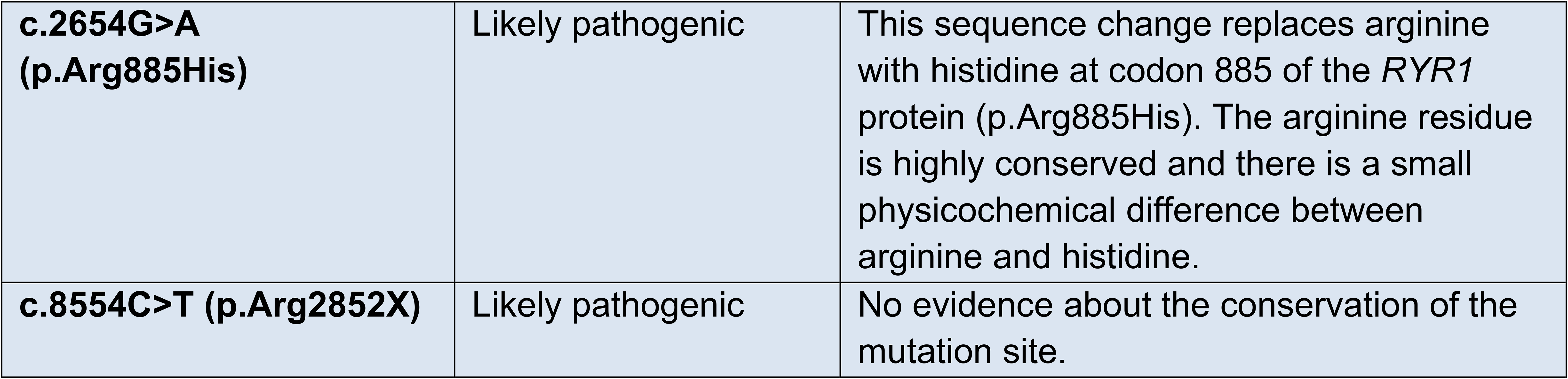
Conservation of mutation sites. Evidence was obtained from Clinvar.

**Online resource table S5:**
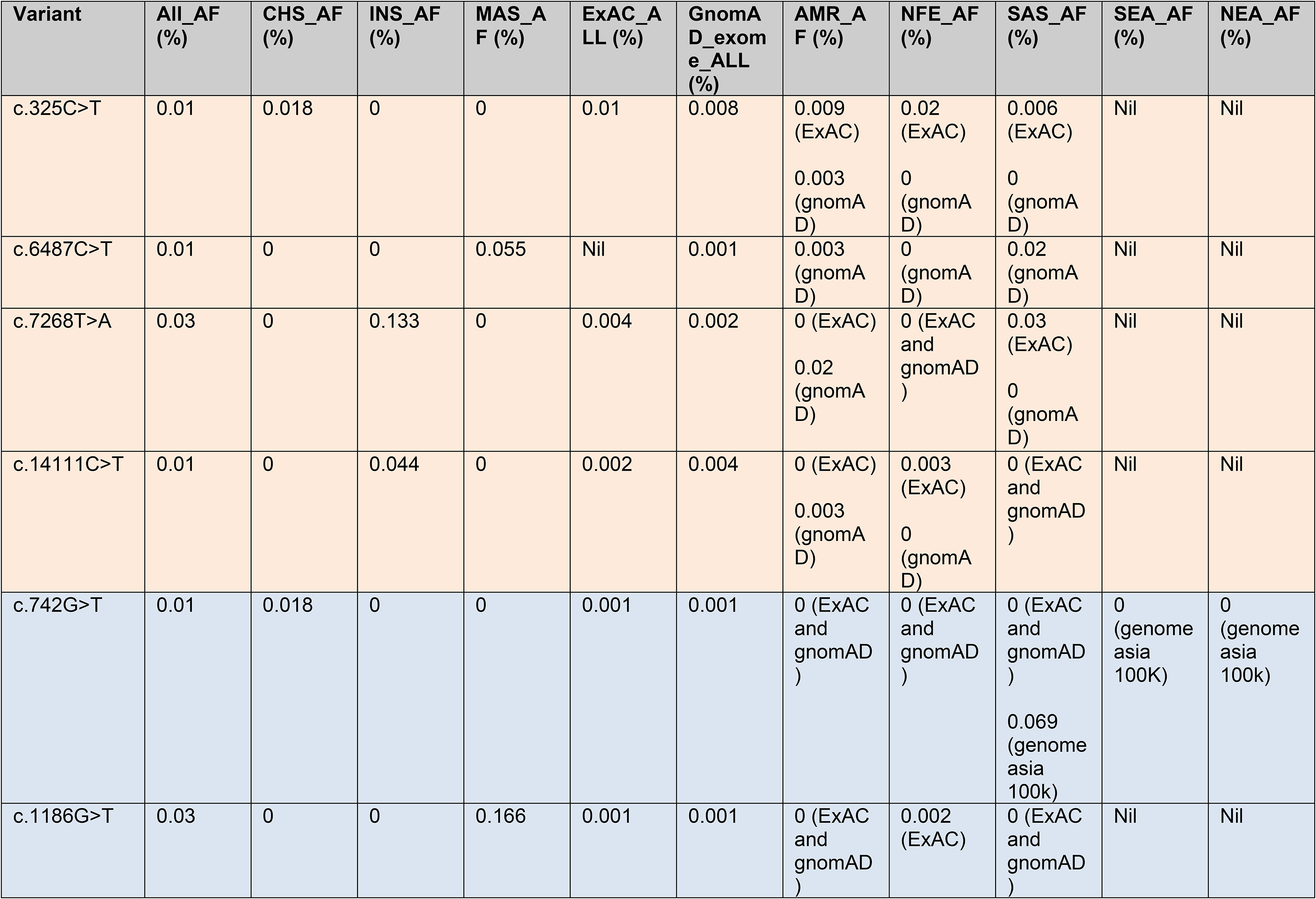
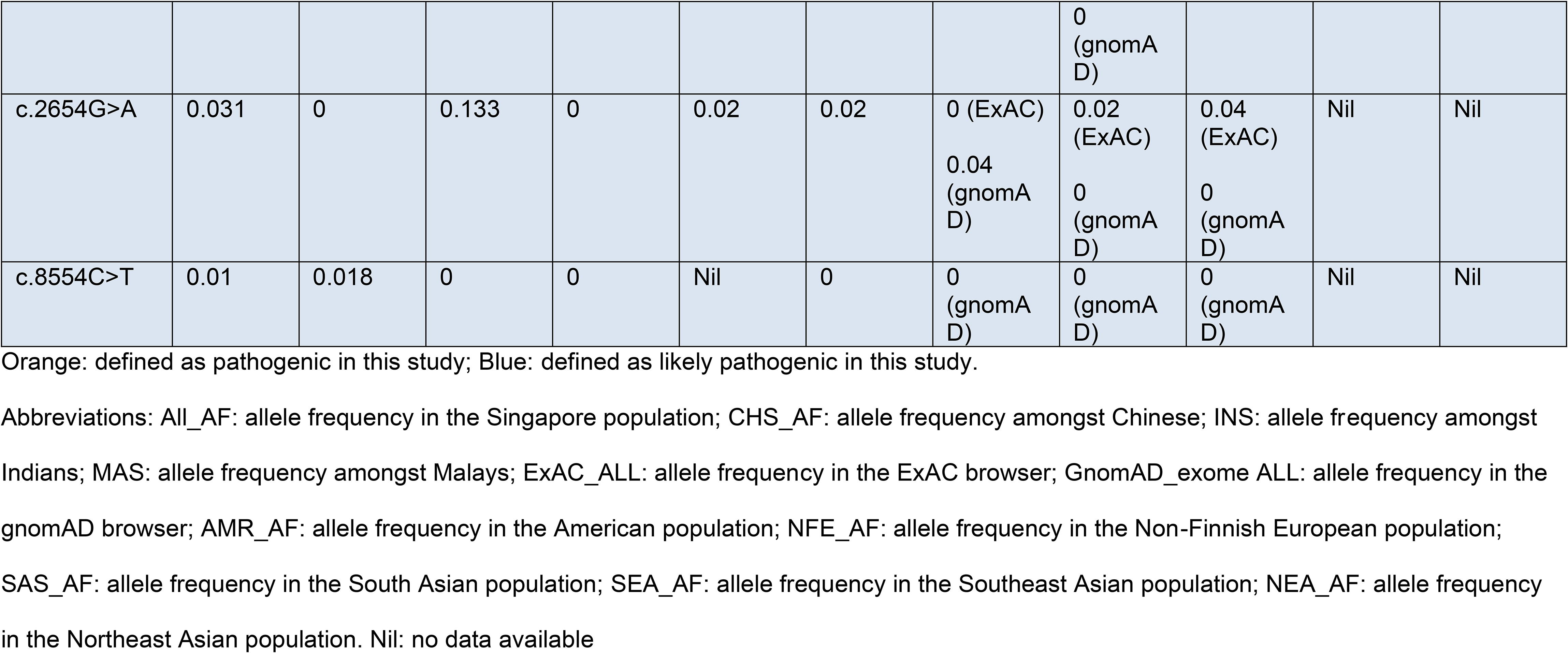
Allele frequencies of the pathogenic and likely pathogenic *RYR1* variants

