## Supplementary figures and images for "Variant landscape of the *RYR1 gene* based on whole genome sequencing of the Singaporean population"

### Online resource figure S1

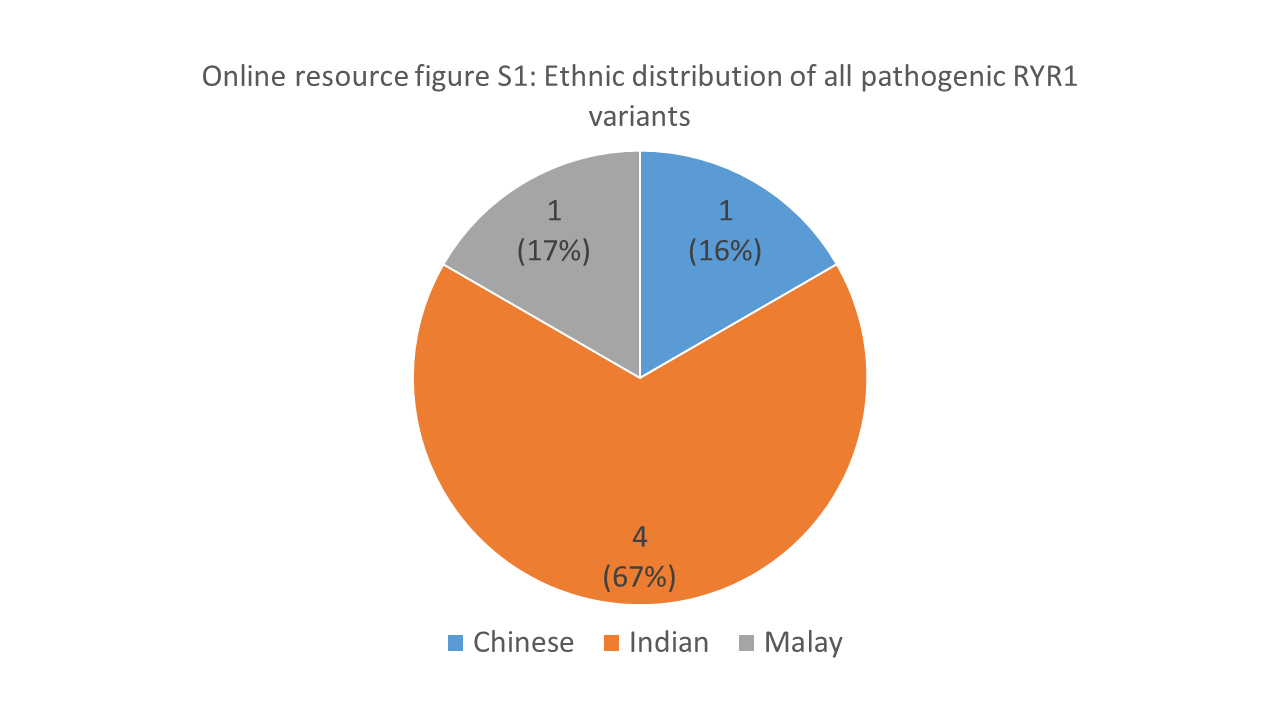

### Online resource figure S2

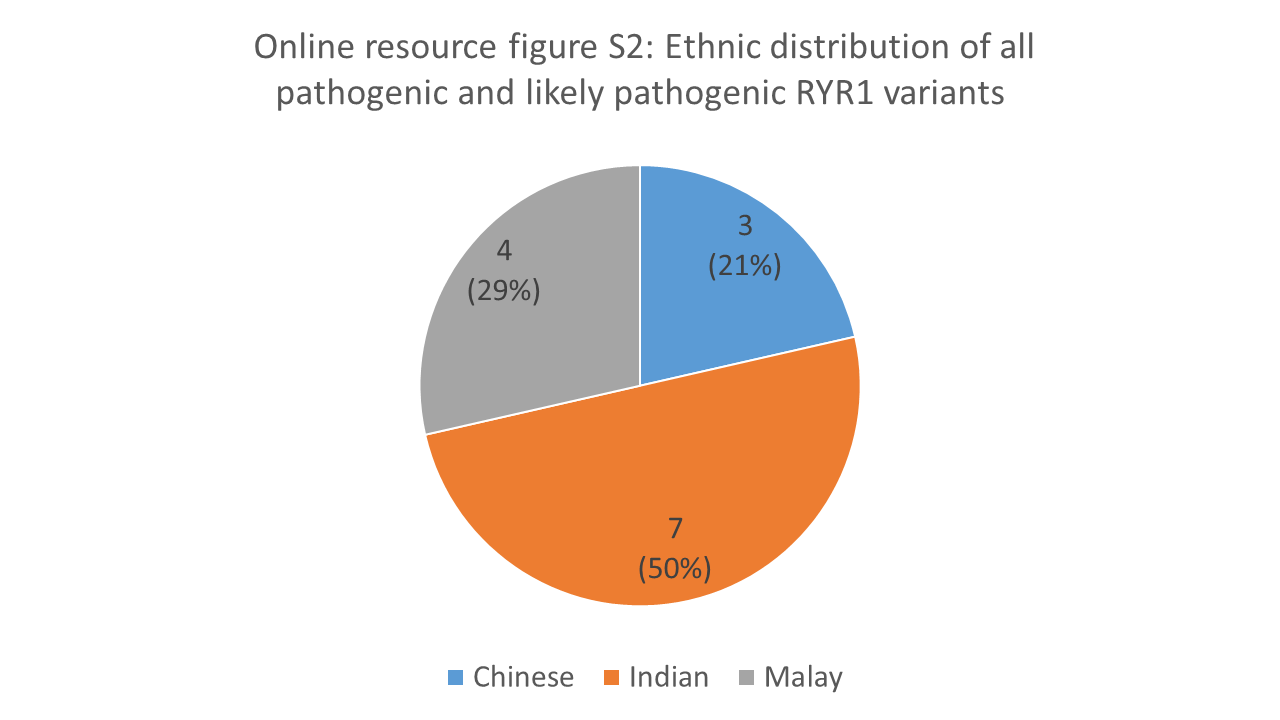

### Online resource figure S3

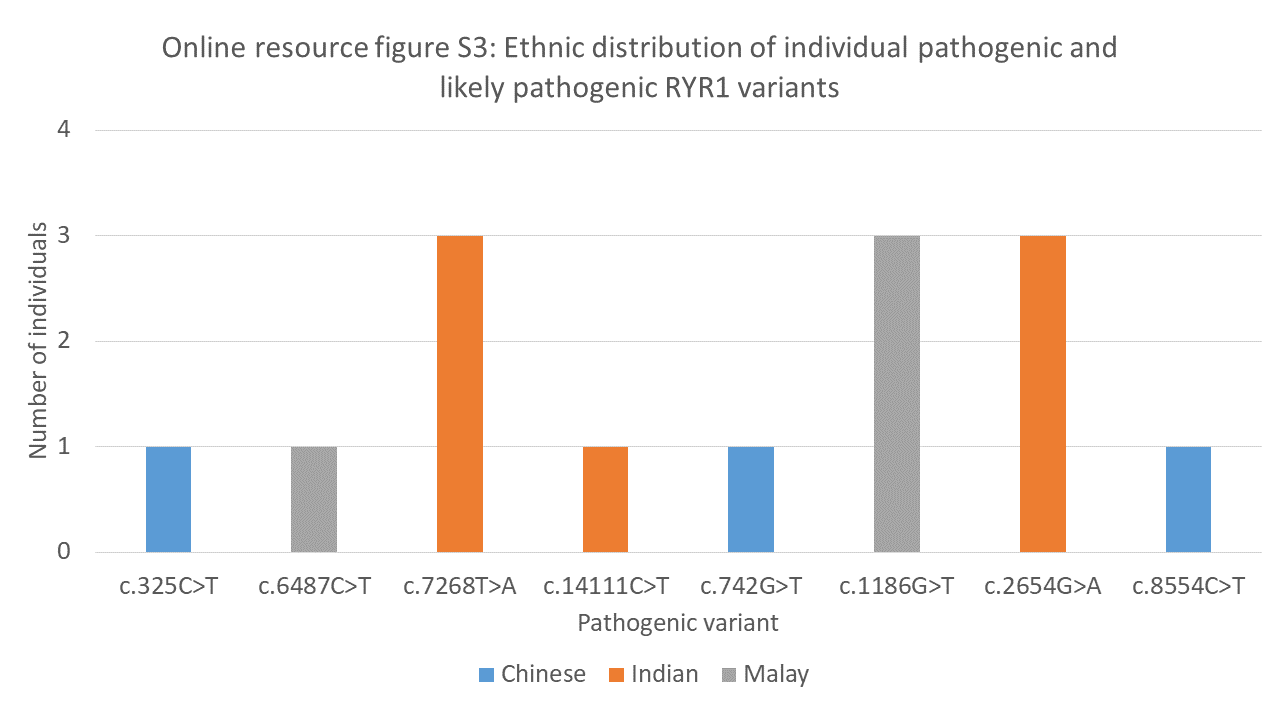
