## Supplementary material for "Variant landscape of the *RYR1 gene* based on whole genome sequencing of the Singaporean population": Online resource table S1

Online resource table S1: Clinical significance of *RYR1* variants based on ClinVar.

| Clinvar classification | Number (percentage) of *RYR1* variants |
|---|---|
| **Pathogenic** | 4 (0.7%) |
| **Likely Pathogenic** | 2 (0.3%) |
| **Variant of uncertain significance** | 88 (15.6%) |
| **Likely benign** | 0 (0.0%) |
| **Benign** | 0 (0.0%) |
| **Conflicting*** | 33 (5.9%) |
| **Unclassified** | 437 (77.5%) |

*Conflicting refers to when different submitters assign different clinical significance to a variant. This includes variants which have the following combination of classifications from different submitters:

1. (Pathogenic or Likely pathogenic or Benign or Likely benign) AND Uncertain significance

2. (Pathogenic or Likely pathogenic) AND (Benign or Likely benign)
