## Supplementary material for "Variant landscape of the *RYR1 gene* based on whole genome sequencing of the Singaporean population": Online resource table S2

Online resource table S2: Clinical significance of *RYR1* variants based on InterVar.

| Intervar classification | Number (percentage) of *RYR1* variants |
|---|---|
| **Pathogenic** | 1 (0.2%) |
| **Likely Pathogenic** | 6 (1.1%) |
| **Variant of uncertain significance** | 274 (48.5%) |
| **Likely benign** | 237 (42.0%) |
| **Benign** | 28 (5.0%) |
| **Unclassified** | 18 (3.2%) |
