## Supplementary material for "Variant landscape of the *RYR1 gene* based on whole genome sequencing of the Singaporean population": Online resource table S3

Online resource table S3: Diseases associated with pathogenic and likely pathogenic RYR1 variants with supporting evidence from ClinVar

| Variant | Interpreted conditions | ClinVar interpretation | Evidence |
|---|---|---|---|
| c.325C>T (p.Arg109Trp) | Minicore myopathy with external opthalmoplegia | Pathogenic | Mutation found in 2 minicore myopathy patients from 1 family with disease onset in the neonatal period (https://pubmed.ncbi.nlm.nih.gov/16380615/) |
| | Congenital myopathy with fibre type disproportion | Likely pathogenic | The following ACMG criteria were applied in classifying this variant: PS1, PM2, PP3*. |
| c.6487C>T (p.Arg2163Cys) | 1. Malignant hyperthermia susceptibility<br>2. Central core myopathy | Pathogenic | 1. Mutation found in individuals from 2 families who had malignant hyperthermia episodes (https://www.cell.com/ajhg/fulltext/S0002-9297(07)63840-3); annotated in PharmGKB database as associated with Malignant Hyperthermia (Level 1A evidence) (https://www.pharmgkb.org/clinicalAnnotation/1183705797)<br>2. Submitted from GeneReviews, converted during submission to pathogenic (https://www.ncbi.nlm.nih.gov/clinvar/submitters/500062/) |
| c.7268T>A (p.Met2423Lys) | 1. Minicore myopathy with external ophthalmoplegia<br>2. Congenital myopathy with fibre type disproportion<br>3. Clubfoot<br>4. EMG abnormality<br>5. Lower limb amyotrophy | Pathogenic | 1. Mutation found in 4 individuals from 1 family with disease onset in infancy and childhood (https://pubmed.ncbi.nlm.nih.gov/16380615/)<br>2-5. Submitted by Centre for Mendelian Genomics, University Medical Centre Ljubljana (https://www.ncbi.nlm.nih.gov/clinvar/submitters/505952/) |
| c.14111C>T (p.Thr4704Met) | 1. Minicore myopathy with | Pathogenic | 1. Mutation associated with clinical features of the disease and molecular studies showed an |

| | | | |
|---|---|---|---|
| | external ophthalmoplegia<br>2. Central core myopathy | | association with lower levels of RYR1 protein on western blot analysis (https://pubmed.ncbi.nlm.nih.gov/17483490/)<br>2. Submitted from GeneReviews (https://www.ncbi.nlm.nih.gov/clinvar/variation/65996/evidence/) |
| | 1. Central core myopathy<br>2. Congenital myopathy with fiber type disproportion<br>3. Malignant hyperthermia susceptibility<br>4. Minicore myopathy with external ophthalmoplegia | Likely pathogenic | Submitted from Fulgent Genetics (https://www.ncbi.nlm.nih.gov/clinvar/variation/65996/evidence/) |
| | Malignant hyperthermia susceptibility | Variant of uncertain significance | This variant was observed as part of a predisposition screen in an ostensibly healthy population. A literature search was performed for the gene, cDNA change, and amino acid change (where applicable). No publications were found based on this search. Allele frequency data from public databases did not allow this variant to be ruled in or out of causing disease. Therefore, this variant is classified as a variant of unknown significance. (https://www.ncbi.nlm.nih.gov/clinvar/variation/65996/evidence/) |
| c.742G>T (p.Gly248Trp) | Malignant hyperthermia | Likely pathogenic | Segregated with malignant hyperthermia in 1 out of 45 families tested (https://www.sciencedirect.com/science/article/abs/pii/088875439290042Q) |
| c.1186G>T (p.Glu396X)** | NA | NA | NA |

| | | | |
|---|---|---|---|
| c.2654G>A (p.Arg885His) | 1. Malignant hyperthermia susceptibility<br>2. Minicore myopathy with external ophthalmoplegia<br>3. Congenital myopathy with fiber type disproportion | Uncertain significance | 1. This variant was observed in the ICSL laboratory as part of a predisposition screen in an ostensibly healthy population. It had not been previously curated by ICSL or reported in the Human Gene Mutation Database (HGMD: prior to June 1st, 2018), and was therefore a candidate for classification through an automated scoring system. Utilizing variant allele frequency, disease prevalence and penetrance estimates, and inheritance mode, an automated score was calculated to assess if this variant is too frequent to cause the disease. Based on the score, this variant could not be ruled out of causing disease and therefore its association with disease required further investigation. A literature search was performed for the gene, cDNA change, and amino acid change (if applicable). No publications were found based on this search. This variant was therefore classified as a variant of unknown significance for this disease. (https://www.ncbi.nlm.nih.gov/clinvar/variation/212100/evidence/)<br>2, 3. Submitted from Genomic Research Center, Shahid Beheshti University of Medical Services and Illumina Clinical Services Laboratory |
| c.8554C>T (p.Arg2852X) | RYR1-related disorders | Variant of uncertain significance | The RYR1 c.8554C>T (p.Arg2852Ter) stop-gained variant has been reported in one study in which it was found in a compound heterozygous state in one individual with muscular dystrophy and arthrogryposis (Vasli et al. 2012). The individual's affected twin brother was a compound heterozygote for the |

| | | | |
|---|---|---|---|
| | | | same two variants, but it is not known whether the twin brothers were monozygotic or dizygotic. The p.Arg2852Ter variant was also found in a heterozygous state in an unaffected sibling and an unaffected parent. Control data are unavailable for this variant, and the variant is not found in the 1000 Genomes Project, the Exome Sequencing Project, or the Exome Aggregation Consortium. The disease description in this family is most consistent with multiminicore disease, but there is considerable overlap of disease symptoms with central core disease and congenital neuromuscular disease with uniform type 1 fiber. **The p.Arg2852Ter variant has not been reported in the literature in association with malignant hyperthermia susceptibility**. Due to the potential impact of stop-gained variants and the evidence from the literature, the p.Arg2852Ter variant is classified as a variant of unknown significance but suspicious for pathogenicity for RYR1-related disorders. This variant was observed by ICSL as part of a predisposition screen in an ostensibly healthy population. (https://www.ncbi.nlm.nih.gov/clinvar/variation/329081/evidence/) |

*PS1: same amino acid change known; PM2: absent from controls; PP3: in silico evidence

**Not found in ClinVar. Labelled as pathogenic in InterVar.
