## Supplementary material for "Variant landscape of the *RYR1 gene* based on whole genome sequencing of the Singaporean population": Online resource table S4

Online resource table S4: Conservation of mutation sites. Evidence was obtained from Clinvar.

| Variant | Clinical significance | Evidence |
|---|---|---|
| **c.325C>T (p.Arg109Trp)** | Pathogenic | This sequence change replaces arginine with tryptophan at codon 109 of the *RYR1* protein (p.Arg109Trp). The arginine residue is highly conserved and there is a moderate physicochemical difference between arginine and tryptophan. |
| **c.6487C>T (p.Arg2163Cys)** | Pathogenic | The R2163C variant is a non-conservative amino acid substitution, which occurs at a position that is conserved across species. |
| **c.7268T>A (p.Met2423Lys)** | Pathogenic | In 3 sibs with minicore myopathy with external ophthalmoplegia (255320) originally reported by Swash and Schwartz (1981), Jungbluth et al. (2005) identified a 7268T-A transversion in exon 45 the *RYR1* gene, resulting in a met2423-to-lys substitution in a highly conserved region. |
| **c.14111C>T (p.Thr4704Met)** | Pathogenic | This sequence change replaces threonine with methionine at codon 4709 of the *RYR1* protein (p.Thr4709Met). The threonine residue is highly conserved and there is a moderate physicochemical difference between threonine and methionine. |
| **c.742G>T (p.Gly248Trp)** | Likely pathogenic | No evidence about the conservation of the mutation site. |
| **c.1186G>T (p.Glu396X)** | Likely pathogenic | No evidence about the conservation of the mutation site. |
| **c.2654G>A (p.Arg885His)** | Likely pathogenic | This sequence change replaces arginine with histidine at codon 885 of the *RYR1* protein (p.Arg885His). The arginine residue is highly conserved and there is a small physicochemical difference between arginine and histidine. |
| **c.8554C>T (p.Arg2852X)** | Likely pathogenic | No evidence about the conservation of the mutation site. |
