## Supplementary material for "Variant landscape of the *RYR1 gene* based on whole genome sequencing of the Singaporean population": Online resource table S5

Online resource table S5: Allele frequencies of the pathogenic and likely pathogenic *RYR1* variants

| **Variant** | **All_AF (%)** | **CHS_AF (%)** | **INS_AF (%)** | **MAS_AF (%)** | **ExAC_ALL (%)** | **GnomAD_exome_ALL (%)** | **AMR_AF (%)** | **NFE_AF (%)** | **SAS_AF (%)** | **SEA_AF (%)** | **NEA_AF (%)** |
|---|---|---|---|---|---|---|---|---|---|---|---|
| c.325C>T | 0.01 | 0.018 | 0 | 0 | 0.01 | 0.008 | 0.009 (ExAC)<br><br>0.003 (gnomAD) | 0.02 (ExAC)<br><br>0 (gnomAD) | 0.006 (ExAC)<br><br>0 (gnomAD) | Nil | Nil |
| c.6487C>T | 0.01 | 0 | 0 | 0.055 | Nil | 0.001 | 0.003 (gnomAD) | 0 (gnomAD) | 0.02 (gnomAD) | Nil | Nil |
| c.7268T>A | 0.03 | 0 | 0.133 | 0 | 0.004 | 0.002 | 0 (ExAC)<br><br>0.02 (gnomAD) | 0 (ExAC and gnomAD) | 0.03 (ExAC)<br><br>0 (gnomAD) | Nil | Nil |
| c.14111C>T | 0.01 | 0 | 0.044 | 0 | 0.002 | 0.004 | 0 (ExAC)<br><br>0.003 (gnomAD) | 0.003 (ExAC)<br><br>0 (gnomAD) | 0 (ExAC and gnomAD) | Nil | Nil |
| c.742G>T | 0.01 | 0.018 | 0 | 0 | 0.001 | 0.001 | 0 (ExAC and gnomAD) | 0 (ExAC and gnomAD) | 0 (ExAC and gnomAD)<br><br>0.069 (genome | 0 (genome asia 100K) | 0 (genome asia 100k) |

| | | | | | | | | | | | |
|---|---|---|---|---|---|---|---|---|---|---|---|
| | | | | | | | | | asia 100k) | | |
| c.1186G>T | 0.03 | 0 | 0 | 0.166 | 0.001 | 0.001 | 0 (ExAC and gnomAD ) | 0.002 (ExAC)<br><br>0 (gnomAD) | 0 (ExAC and gnomAD ) | Nil | Nil |
| c.2654G>A | 0.031 | 0 | 0.133 | 0 | 0.02 | 0.02 | 0 (ExAC)<br><br>0.04 (gnomAD) | 0.02 (ExAC)<br><br>0 (gnomAD) | 0.04 (ExAC)<br><br>0 (gnomAD) | Nil | Nil |
| c.8554C>T | 0.01 | 0.018 | 0 | 0 | Nil | 0 | 0 (gnomAD) | 0 (gnomAD) | 0 (gnomAD) | Nil | Nil |

Orange: defined as pathogenic in this study; Blue: defined as likely pathogenic in this study.

Abbreviations: All_AF: allele frequency in the Singapore population; CHS_AF: allele frequency amongst Chinese; INS: allele frequency amongst Indians; MAS: allele frequency amongst Malays; ExAC_ALL: allele frequency in the ExAC browser; GnomAD_exome ALL: allele frequency in the gnomAD browser; AMR_AF: allele frequency in the American population; NFE_AF: allele frequency in the Non-Finnish European population; SAS_AF: allele frequency in the South Asian population; SEA_AF: allele frequency in the Southeast Asian population; NEA_AF: allele frequency in the Northeast Asian population. Nil: no data available
